# Mechanical Thrombectomy for Medium and Distal Vessel Occlusion: A Systematic Review and Meta-Analysis

**DOI:** 10.64898/2026.01.09.26343822

**Authors:** Victória Luiza Macedo Miranda Baldim, João Vitor da Cunha Costa, Layla Moreira Batista, Julia Alves Banzati Viana, Thaíse Uchôas Gonçalves, Rodrigo Corrêa Falcão Rodrigues Alves

**Affiliations:** Medical Student, Faculty of Medical Sciences of São José dos Campos – Humanitas

**Keywords:** Ischemic Stroke, Mechanical Trhombectomy, Intracranial Arterial Occlusion

## Abstract

**Objective:** We aimed to do an updated meta- analysis comparing outcomes of mechanical thrombectomy (MT) associated with standard medical treatment (SMT), compared to SMT alone, in adult patients with acute ischemic stroke (AIS) due to medium or distal vessel occlusion (MDVO).

**Methodology:** We systematically searched PubMed, LILACS, Scielo, Cochrane, and ClinicalTrials.gov databases. Randomized controlled trials (RCTs), retrospective cohort studies, and systematic reviews with meta-analysis comparing MT+SMT with SMT alone in adults with AIS due to MDVO, evaluating at least one of the outcomes of interest, were included. The evaluated outcomes were functional recovery (modified Rankin Scale [mRS] 0-1 and mRS 0-2) at 90 days, all-cause mortality at 90 days, and the occurrence of intracranial hemorrhage (ICH). Risk of bias was assessed using RoB 2, Newcastle-Ottawa, and AMSTAR 2 tools. Heterogeneity was assessed with Chi² and I², and publication bias with funnel plots and Egger/Begg tests.

**Results:** Twelve studies (4 RCTs, 7 cohorts, 1 systematic review) were included. Meta-analyses showed no significant difference between MT+SMT and SMT alone for: mRS 0-1 (Excellent Recovery), mRS 0-2 (Good Recovery), mortality at 90 days, and Symptomatic Intracranial Hemorrhage.

**Conclusion:** Current evidence, combining RCTs and observational studies, does not support the routine use of MT over SMT alone for MDVO in terms of functional improvement at 90 days (mRS 0-1 or 0-2). Non-significant trends towards increased mortality and sICH risk with MT were observed, with considerable heterogeneity for sICH.

**KEY MESSAGE:** Several randomized controlled trials (RCTs) have demonstrated the benefit of mechanical thrombectomy associated with standard medical treatment (SMT), in patients with IS caused by large vessel occlusion (LVO). There for the boundaries of MT began to be questioned, raising the possibility of performing it for medium and distal vessel occlusions (MDVO). As a result of this study, there was no evidence to support the use of MT over SMT alone for MDVO, therefore the therapeutic decisions should remain individualized and further research is crucial to define the exact role of the MT.

## INTRODUCTION

Ischemic Stroke (IS) represents one of the leading causes of mortality and disability globally ^(1)^. In recent decades, the treatment of IS has undergone significant transformations, culminating in the advent and implementation of mechanical thrombectomy (MT). Several randomized controlled trials (RCTs), published mainly since 2015, have demonstrated the benefit of MT associated with standard medical treatment (SMT), in patients with IS caused by large vessel occlusion (LVO) ^(2–6)^. The HERMES collaboration meta-analysis confirmed the robustness of this benefit, establishing MT as the gold standard for these patients ^(7)^.

Consequently, the boundaries of MT began to be questioned, raising the possibility of performing it for medium and distal vessel occlusions (MDVO), generally defined as M2 and M3 segments of the middle cerebral artery (MCA), A2 and A3 segments of the anterior cerebral artery (ACA), and P1-P3 segments of the posterior cerebral artery (PCA) (4–9). Such occlusions, although often associated with initially less severe neurological deficits as measured by the National Institutes of Health Stroke Scale (NIHSS), can result in significant long-term disabilities ^(8–10)^. Observational studies initially suggested potential benefits of MT for MDVO, particularly for M2 occlusions ^(11–13)^. However, the level of evidence remained low, based mainly on retrospective cohort studies with significant methodological limitations, such as selection bias and confounding factors.

Recently, the results of the first RCTs specifically designed to evaluate MT in MDVO were published: ESCAPE-MeVO ^(14)^ and DISTAL ^(15)^. Both trials failed to demonstrate a significant benefit of MT compared to SMT alone regarding functional outcomes at 90 days. ESCAPE-MeVO even suggested a trend towards harm, with lower rates of excellent functional outcome (mRS 0-1) in the MT group. The DISTAL trial showed numerically better outcomes for SMT, although not statistically significant. These results contrast with previous observational evidence and raise questions about the real efficacy and safety of MT in this specific subgroup of IS patients. Preliminary results from other ongoing trials, such as DISCOUNT ^(16)^, also seem to align with these findings.

Given the conflicting results between observational studies and recent RCTs, and the clinical importance of defining the best therapeutic strategy for MDVO, a comprehensive and updated synthesis of the available evidence is crucial. Therefore, our review aims to critically evaluate the efficacy and safety of MT associated with SMT, compared to SMT alone, in adult patients with acute IS due to MDVO. We evaluated functional outcomes at 90 days (mRS 0-1 and mRS 0-2), mortality at 90 days, and the occurrence of intracranial hemorrhage (ICH).

## METHODOLOGY

This is a systematic literature review with meta-analysis on the performance of mechanical thrombectomy (MT) for ischemic stroke (IS) of medium and distal vessels (MDVO) in contrast to isolated standard medical therapy (SMT). A research protocol was written primarily. Our study follows the guidelines proposed by the Preferred Reporting Items for Systematic Reviews and Meta-Analyses (PRISMA) and is registered on the PROSPERO platform (PROSPERO 2025 CRD420251035010). Two authors systematically searched for retrospective cohort studies, randomized clinical trials, and systematic reviews on mechanical thrombectomy in patients with ischemic stroke, within the period from January 2015 to April 2025 in the PubMed, LILACS, Scielo, Cochrane, and ClinicalTrials.gov databases. The terms used were *ischemic stroke AND thrombectomy AND medium vessel*. As outcomes, the modified Rankin score (mRS) between 0-1 at 90 days, mRS between 0-2 at 90 days, mortality at 90 days, and occurrence of intracranial hemorrhage were primarily evaluated.

### Eligibility Criteria

Studies meeting the following criteria (PICOS) were included:

- **Population:** adult patients (age > 18 years), with IS due to medium or distal vessel occlusion.
- **Intervention:** mechanical thrombectomy associated with standard medical treatment.
- **Comparator:** standard medical treatment alone.
- **Outcomes:** at least one of the following: (1) mRS 0-1 at 90 days – excellent functional outcome; (2) mRS 0-2 at 90 days – good/favorable functional outcome;
- (3) all-cause mortality at 90 days; (4) intracranial hemorrhage.
- **Study types:** randomized clinical trials, retrospective cohort studies, and systematic reviews with meta-analysis.

After the search, two authors selected studies based on the title and abstract of the articles and according to the established inclusion and exclusion criteria, as shown in Figure 1. Conflicts were resolved by a third author.

**Fig. 1:**
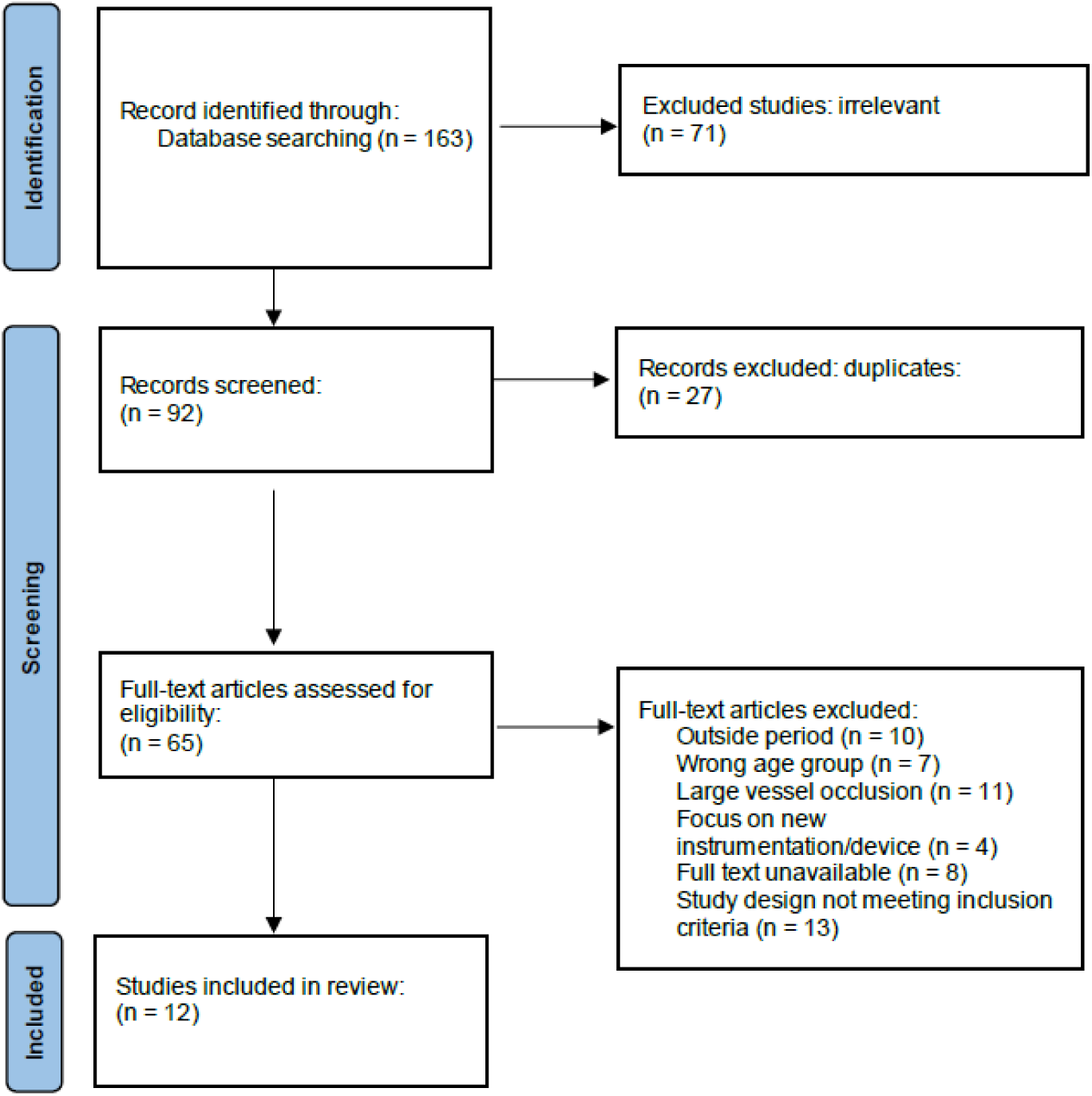
Flowchart of included article selection. Adapted from Preferred Reporting Items for Systematic Reviews and Meta-Analyses (PRISMA).

**Fig. 2:**
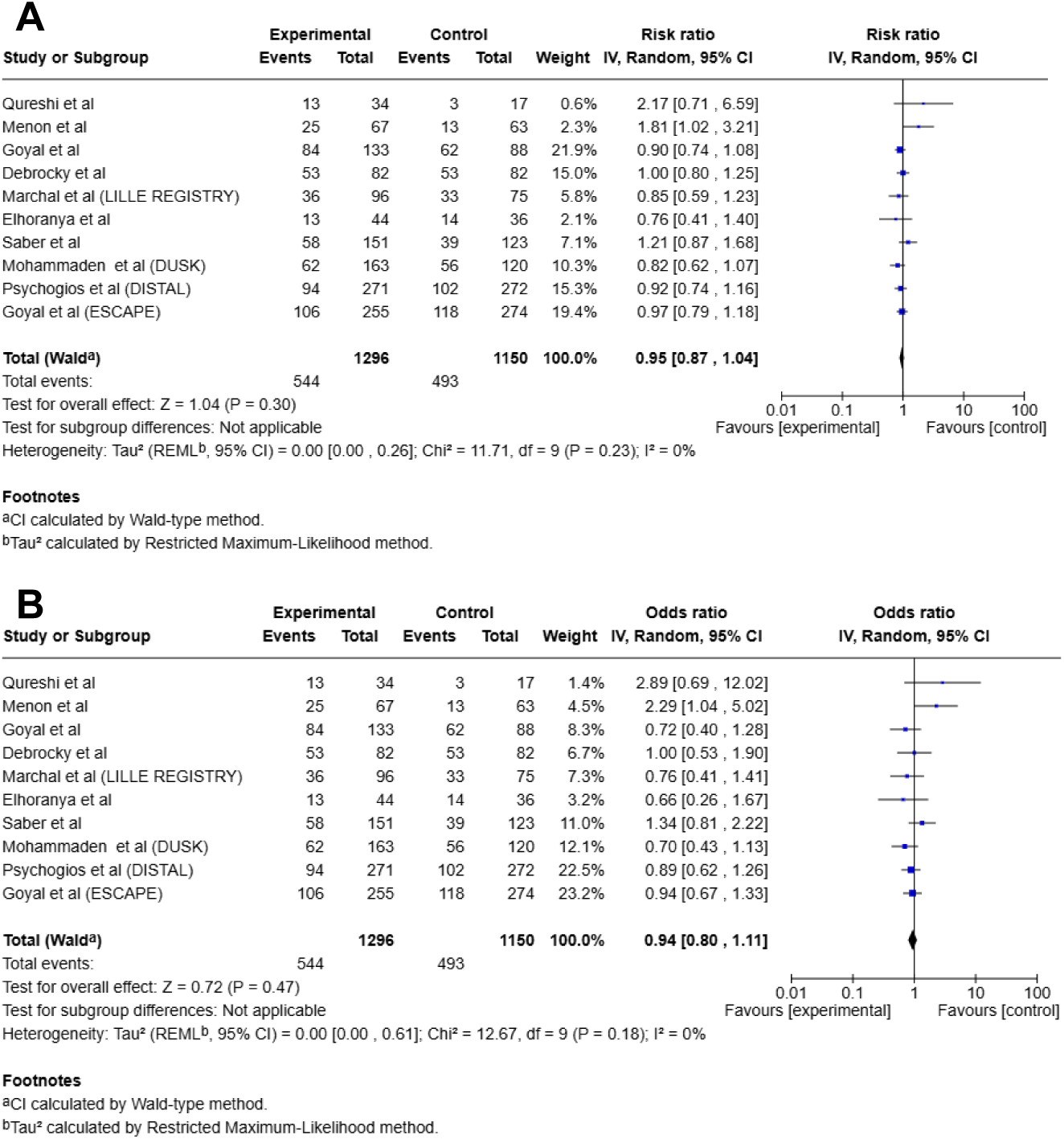
Forest plots comparing the mRS 0-1 outcome between TM+TC (experimental) with TC alone (control). (A) represents Risk Ratio and (B), Odds Ratio.

**Fig. 3:**
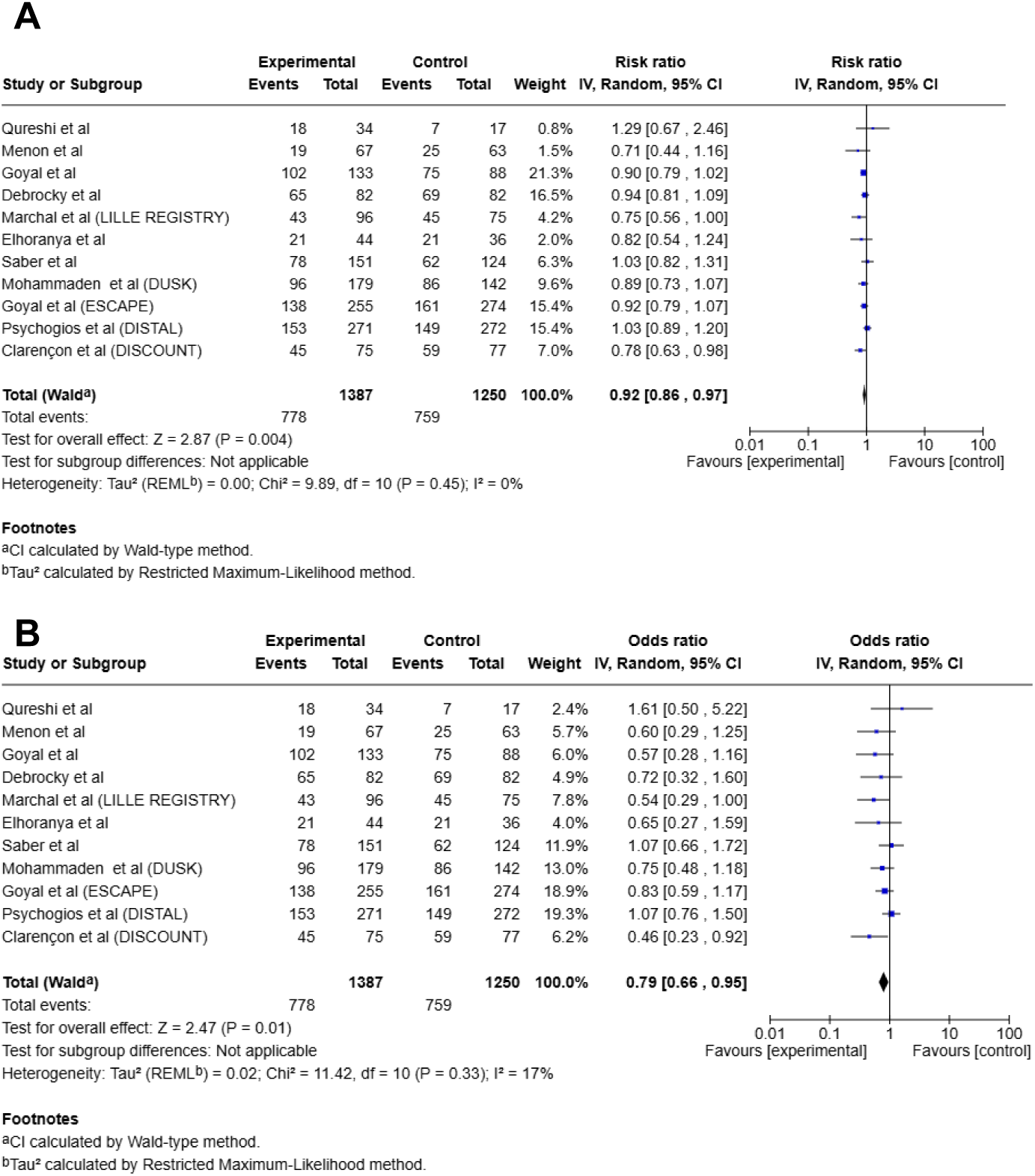
Forest plots comparing the mRS 0-2 outcome between TM+TC (experimental) with TC alone (control). (A) represents Risk Ratio and (B), Odds Ratio.

**Fig. 4:**
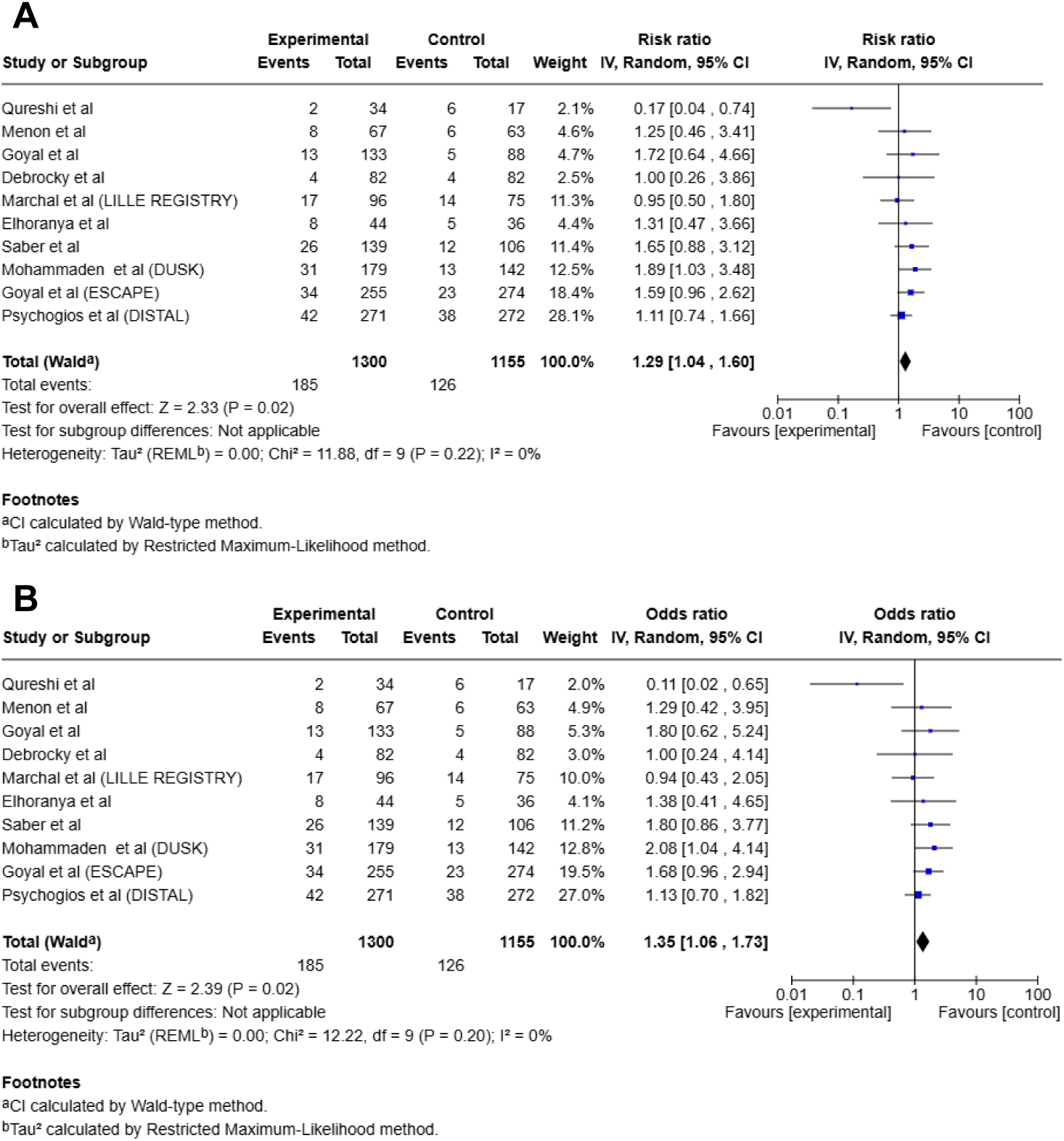
Forest plots comparing the 90-day mortality outcome between TM+TC (experimental) and TC alone (control). (A) represents Risk Ratio and (B), Odds Ratio.

**Fig. 5:**
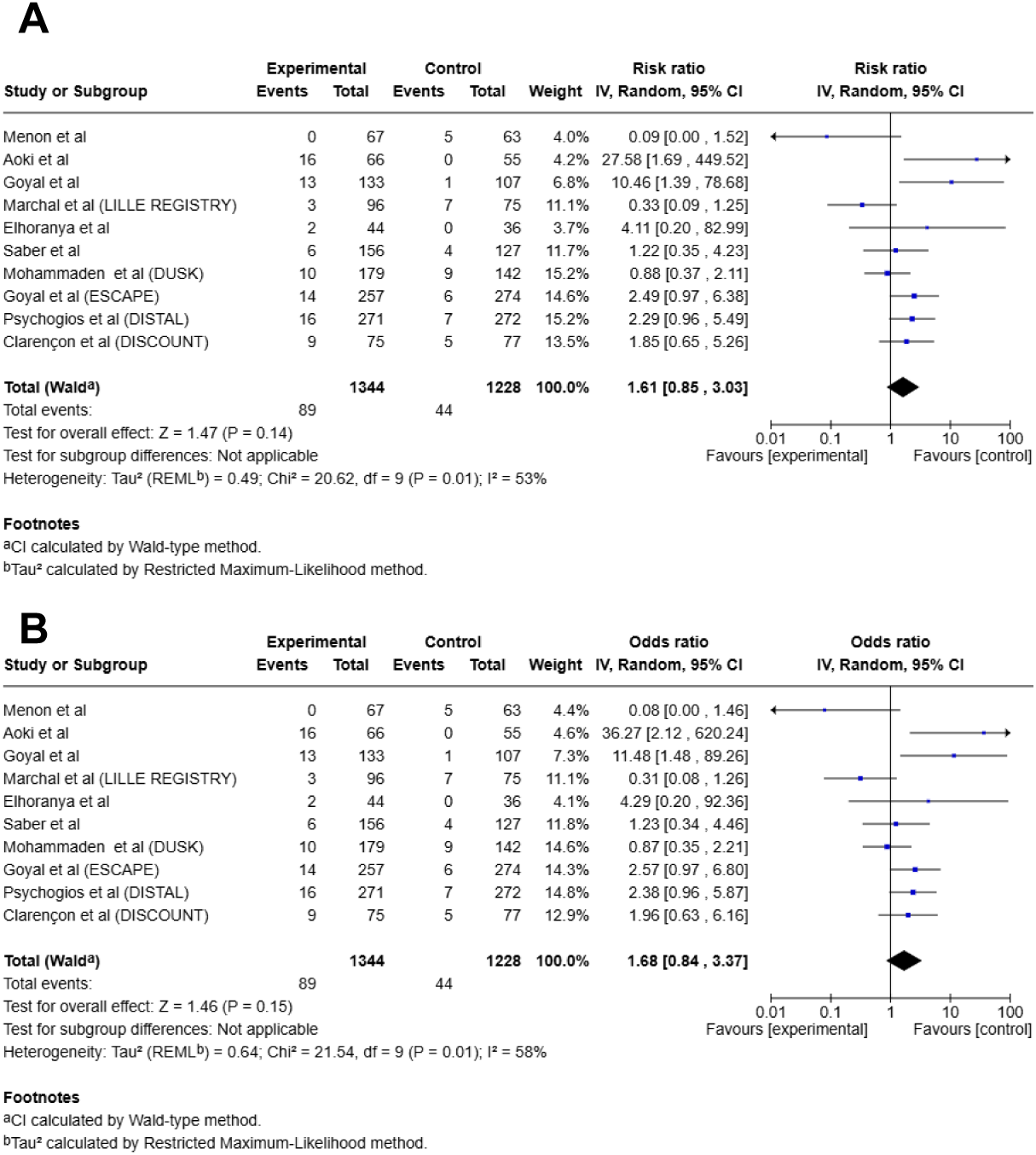
Forest plots comparing the outcome of intracranial hemorrhage between MT+CT (experimental) and CT alone (control). (A) represents Risk Ratio and (B), Odds Ratio.

**Inclusion criteria:** studies from the last 10 years, medium and distal vessel occlusion, randomized clinical trials, retrospective cohorts, adults.

**Exclusion criteria:** involved studies with large vessel occlusion, non-human studies, duplicate studies, studies comparing and/or presenting new thrombectomy techniques, studies presenting new instrumentation for thrombectomy, studies containing patients with IS plus other neurological diseases.

In the end, a total of 12 studies were analyzed, four randomized clinical trials, seven retrospective cohorts and one systematic review with meta-analysis.

Data extraction from the included studies was performed independently by two reviewers, using a standardized data extraction form, including year, first author, design, number of participants, population characteristics (age, sex), and NIHSS (Table 1). The extractions were then compared, and any discrepancies were resolved by consensus between the two reviewers or, when necessary, with consultation from a third reviewer. For the selected studies, the full articles were read.

**Table 1:** Standardized data collection.

| Study | Goyal et al (2025) | Psychogios et al (2025) | Mohammaden et al (2024) | Marchal et al (2022) | Clarençon et al (2025) | Aoki et al (2020) |
| --- | --- | --- | --- | --- | --- | --- |
| Design | ECR | Cohort | Cohort | Cohort | ECR | Cohort |
| **Thrombectomy + Clinical treatment** |  |  |  |  |  |  |
| Participants | 255 | 271 | 179 | 96 | 75 | 66 |
| Age (Median) [Mean] | (74) | (77) | [69.4] | [68.7] | - | - |
| Female | 118 | 116 | 118 | 51 | - | - |
| NIHSS Median | 8 | 6 | 10 | 11 | - | - |
| **Clinical treatment alone** |  |  |  |  |  |  |
| Participants | 274 | 272 | 142 | 75 | 77 | 55 |
| Age (Median) [Mean] | (76) | (77.5) | [66.4] | [73.9] | - | - |
| Female | 127 | 123 | 84 | 42 | - | - |
| NIHSS Median | 7 | 6 | 6 | 10 | - | - |
| Study | Debrocky et al (2021) | Elhoranya et al (2022) | Goyal et al (2020) | Menon et al (2019) | Qureshi et al (2017) | Saber et al (2022) |
| Design | Cohort | Cohort | Cohort | Systematic review | ECR | Cohort |
| **Thrombectomy + Clinical treatment** |  |  |  |  |  |  |
| Participants | 85 | 44 | 138 | 67 | 34 | 156 |
| Age (Median) [Mean] | (71.1) | (78) | [65.2] | (66.4) | (71) | [66.7] |
| Female | 33 | 23 | 65 | 36 | 18 | 66 |
| NIHSS Median | 3 | 11 | 4 | 14 | 16 | 13 |
| **Clinical treatment alone** |  |  |  |  |  |  |
| Participants | 84 | 36 | 113 | 64 | 17 | 130 |
| Age (Median) [Mean] | (67.7) | (70) | [64.8] | (65.9) | (73) | [69.8] |
| Female | 32 | 15 | 51 | 30 | 5 | 68 |
| NIHSS Median | 3 | 6 | 3 | 15 | 14 | 7 |

Randomized clinical trials were evaluated using the RoB 2 tool (Revised Cochrane risk-of-bias tool for randomized trials), which assesses five main domains. Cohort studies were evaluated with the Newcastle-Ottawa tool, which uses three domains and a maximum score of up to nine. Systematic reviews were evaluated by AMSTAR 2. Appendix 1 shows the risk of bias for each study. Funnel plots and tests are reported in the supplementary material.

The assessment of publication bias was performed using a combined approach. Firstly, visual inspection of the symmetry of the funnel plots was carried out for each meta-analysis. In addition, formal statistical tests were applied to assess funnel plot asymmetry: Egger’s regression test and Begg’s rank correlation test. The results indicated that the funnel plots for the four main outcomes appeared reasonably symmetrical upon visual inspection. Furthermore, the results of the Egger (P > 0.14 for all outcomes) and Begg (P > 0.24 for all outcomes) tests did not provide statistical evidence of significant asymmetry for any of the meta-analyses performed. We recognize the inherent limitations of these tests, particularly when the number of studies included in the meta-analysis is relatively small.

The extracted data were combined by meta-analysis for each of the four outcomes. The effect measure used was Odds Ratio (OR) and Relative Risk (RR) with 95% Confidence Intervals (95% CI), as presented in the generated Forest Plots. For all analyses, a random-effects model was used, considering potential clinical and methodological heterogeneity. Statistical heterogeneity among studies was assessed using the Chi-square (Chi2) test and quantified by the I² statistic. The interpretation of I² followed Cochrane recommendations: 0% to 40% (low or unimportant heterogeneity); 30% to 60% (may represent moderate heterogeneity); 50% to 90% (may represent substantial heterogeneity); 75% to 100% (considerable heterogeneity). A p-value < 0.10 for the Chi2 test was considered indicative of statistically significant heterogeneity. Assessment of publication bias was done using Egger and Begg tests (supplementary material). RevMan and MedCalc software were used for statistical analyses.

## RESULTS

After the systematic search in the databases and application of eligibility criteria, 12 studies were included in this systematic review and meta-analysis, as detailed in the PRISMA flowchart (Figure 1). The included studies consisted of four randomized clinical trials (RCTs), seven cohort studies, and one systematic review with meta-analysis, published between 2017 and 2025. The detailed characteristics of the studies, including design, number of participants, age, sex, and baseline NIHSS, are summarized in Table 1. In total, the outcome analyses included up to 1387 patients in the intervention group (Mechanical Thrombectomy + Medical Treatment) and 1250 in the control group (Medical Treatment alone), depending on the analyzed outcome.

The age of the participants varied among the studies. The median or mean age reported in the MT+SMT groups was generally between 65 and 78 years. In the SMT groups, the median or mean age varied between approximately 65 and 77.5 years. Regarding sex distribution, the overall population was relatively balanced, with approximately 49.4% of participants being female. In the MT+SMT groups, the proportion of women varied among studies (approximately 39% to 66%), resulting in about 52.0% of the total. In the SMT groups, this proportion varied similarly (approximately 29% to 59%), totaling about 46.5%.

The initial neurological severity, assessed by the median NIHSS (National Institutes of Health Stroke Scale) score on admission, showed considerable variation among studies and, in some cases, between groups within the same study. In the MT+SMT groups, the median NIHSS ranged from 3 to 16. In the SMT groups, the median NIHSS ranged from 3 to 15.

### Excellent Functional Recovery (mRS 0-1 at 90 days)

Ten studies contributed to this analysis. The meta-analysis showed no statistically significant difference between the Mechanical Thrombectomy (MT) group and the Standard Medical Treatment (SMT) group. The combined Relative Risk (RR) was 0.96 (95% CI [0.87, 1.07], P=0.479) and the combined Odds Ratio (OR) was 0.96 (95% CI [0.77, 1.18], P=0.664). Heterogeneity among studies was considered low to moderate (I² = 24% for RR, I² = 29% for OR). There was no evidence of significant publication bias (Egger and Begg Tests P>0.1 for both measures).

### Good Functional Recovery (mRS 0-2 at 90 days)

Eleven studies were included in this analysis. No statistically significant difference was found between the groups, although a trend was observed. The combined RR was 0.93 (95% CI [0.86, 1.01], P=0.079), suggesting a marginal trend favoring the SMT group, but without reaching statistical significance. The combined OR was 0.85 (95% CI [0.67, 1.06], P=0.148), also without statistical significance and with a trend favoring SMT. Heterogeneity was moderate (I² = 34% for RR, I² = 42% for OR). There was no evidence of significant publication bias (Egger and Begg Tests P>0.6 for both measures).

### 90-days mortality

Ten studies (with available mortality data) were included. There was no statistically significant difference in mortality between the MT and SMT groups. The combined RR was 1.33 (95% CI [0.96, 1.84], P=0.082), indicating a non-significant trend of higher mortality in the MT group. The combined OR was 1.39 (95% CI [0.96, 2.02], P=0.083), also without statistical significance, but with a similar trend. Heterogeneity was moderate (I² = 44-45%, P≈0.06). There was no evidence of significant publication bias (Egger and Begg Tests P>0.2).

### Symptomatic Intracranial Hemorrhage (sICH)

Ten studies contributed to this analysis. No statistically significant difference was found in the risk of sICH between the MT and SMT groups. The combined RR was 1.62 (95% CI [0.83, 3.14], P=0.155), suggesting a non-significant trend of higher risk with MT. The combined OR was 1.68 (95% CI [0.83, 3.41], P=0.150), with a similar trend. Heterogeneity among studies was statistically significant and substantial (I² = 57-59%, P<0.013 for both measures), indicating considerable variability in the risk of sICH among the included studies. There was no evidence of significant publication bias (Egger and Begg Tests P>0.7).

### Sensitivity Analyses

The meta-analyses were conducted using both Relative Risk (RR) and Odds Ratio (OR) for all outcomes (mRS 0-1, mRS 0-2, mortality at 90 days, sICH). The results showed consistency in the direction of the effect and in the conclusion regarding statistical significance between RR and OR for the mRS 0-1, mortality, and sICH outcomes. For the mRS 0-2 outcome, both measures suggested a trend favoring the control group, without reaching statistical significance (P=0.079 for RR; P=0.148 for OR) in the presented numerical results. This consistency between different metrics reinforces the stability of the main conclusions.

Statistical heterogeneity among studies was quantified using the I² statistic. For the outcomes mRS 0-1 (I²≈24-29%), mRS 0-2 (I²≈34-42%), and Mortality (I²≈44-45%), heterogeneity was considered low to moderate, suggesting relative consistency among studies. However, for the sICH outcome, heterogeneity was substantial and statistically significant (I²≈57-59%, P<0.013), indicating that the effect of MT on the risk of sICH may vary significantly among the included studies, limiting the generalization of the combined result.

For all four analyzed outcomes (mRS 0-1, mRS 0-2, mortality, sICH), both visual inspection of the funnel plots and formal statistical tests (Egger and Begg Tests) detected no evidence of significant bias (all P > 0.1). This suggests that the combined results were likely not substantially distorted.

## DISCUSSION

This systematic review and meta-analysis evaluated the efficacy and safety of mechanical thrombectomy (MT), added to standard medical treatment (SMT), compared to SMT alone for patients with acute ischemic stroke (IS) caused by medium or distal vessel occlusion (MDVO). Our main findings indicate that, based on the combined evidence from 12 studies, MT did not demonstrate a statistically significant benefit in terms of excellent (mRS 0-1) or good (mRS 0-2) functional improvement at 90 days, compared to SMT alone. Furthermore, no significant differences were found in the safety outcomes of mortality at 90 days and symptomatic intracranial hemorrhage (sICH), although non-significant trends towards increased risks with MT were observed for both.

These results markedly contrast with the well-established benefit of MT for large vessel occlusions (LVO) in the anterior circulation, as demonstrated in multiple pivotal randomized clinical trials published in the New England Journal of Medicine (MR CLEAN ^(2)^, ESCAPE ^(3)^, EXTEND-IA ^(4)^, SWIFT PRIME ^(5)^, REVASCAT ^(6)^) and consolidated by the HERMES meta-analysis, published in The Lancet ^(7)^.

The absence of a clear benefit of MT in MDVO may be multifactorial. Patients with MDVO may present with smaller infarcts, have a better natural prognosis, or respond better to isolated medical treatment, including intravenous thrombolysis, whose recanalization rates may be higher in more distal vessels ^(8)^. This potentially reduces the margin for demonstrating an additional benefit of MT compared to LVO, where the prognosis without recanalization is often poor.

Our findings align with recent results from some RCTs focused specifically on MDVO, such as DISTAL ^(10)^ and ESCAPE-MeVO ^(22)^, which also failed to demonstrate a clear benefit of MT for primary functional outcomes. Results from observational studies, such as DUSK ^(11)^, Marchal ^(16)^, and Saber ^(18)^, were also mixed or inconclusive regarding the superiority of MT, reflecting the uncertainty present in the literature ^(11,17–19)^. The post-hoc analysis of the HERMES ^(17)^ collaboration suggested efficacy for M2 occlusions, but with caveats regarding generalization, given selection bias. Our meta-analysis, by including more recent and diverse studies, reinforces the lack of robust evidence of benefit.

The non-significant trend towards higher mortality (RR 1.33 [0.96, 1.84]) and sICH (RR 1.62 [0.83, 3.14]) with MT is noteworthy. Although not statistically significant, these safety signals align with theoretical concerns about the technical challenges and inherent risks of navigation and intervention in smaller caliber, more tortuous, and distal vessels ^(8,16)^. The significant heterogeneity found specifically in the sICH analysis (I²≈57-59%) is an important finding, suggesting that the hemorrhagic risk associated with MT in MDVO may vary substantially depending on patient, technique, or center factors, limiting confidence in the combined effect and highlighting the need for caution.

The strength of this review lies in the systematic approach (PRISMA), the inclusion of the latest data from RCTs and cohorts, covering a considerable number of patients, and the formal assessment of heterogeneity and publication bias. From a clinical perspective, our results reinforce the need for caution in indicating MT for MDVO. In the absence of proven functional benefit and with potential safety signals (trend towards higher mortality and sICH, with heterogeneity for sICH), the decision must be highly individualized. Current AHA/ASA ^(20)^ and ESO ^(21)^ guidelines already reflect this uncertainty, with weak or absent recommendations for MT in many distal segments. Patient selection should carefully consider the clinical deficit (especially if disabling despite low NIHSS), occlusion location (more proximal vs. more distal segments), vascular anatomy, presence of penumbra, and the inherent risks of the procedure for each patient.

For the future, more high-quality RCTs are needed, perhaps focused on specific subgroups of MDVO (e.g., dominant proximal M2 vs. M3, occlusions in eloquent areas) and using outcomes that better capture subtle disability, such as neurocognitive and quality of life assessments, in addition to mRS. Standardization of the definition of MDVO and distal MT techniques would also be beneficial to reduce heterogeneity and allow for more robust comparisons.

### Methodological Quality of RCTs

Two RCTs ^(10,22)^ were classified as having Low overall Risk of bias. All five assessed domains (D1: Bias arising from the randomization process; D2: Bias due to deviations from intended interventions; D3: Bias due to missing outcome data; D4: Bias in measurement of the outcome; D5: Bias in selection of the reported result) were considered low risk for these two studies, contributing positively to the overall robustness of the meta-analysis. One study ^(26)^ was classified as having High overall Risk of bias. Although domains D1, D2, and D4 were assessed as low risk, "Some Concerns" were identified in domain D3 and "High Risk" in domain D5, which may limit confidence in its results and its weight in the combined evidence. Lastly, the trial by Clarençon et al ^(25)^ had inconclusive bias, as the data were provided in the context of an interim/provisional analysis. RCTs with low risk of bias are a strong point, suggesting methodological soundness. Considering studies with high risk and without results, potential biases may occur, partially attenuating confidence in the conclusions.

### Limitations

The inclusion of observational studies (cohorts) and a systematic review alongside RCTs increases the risk of residual confounding bias, despite the use of random-effects models. The definition of MDVO may vary between studies ^(4–9)^, as well as the initial severity of patients (median NIHSS ranged from 3 to 16 in MT groups), contributing to clinical heterogeneity. The evolution of MT techniques and devices over the period covered by the studies may also influence the results. The analysis was based on aggregated data, limiting the exploration of specific subgroups (e.g., by exact occlusion location, eloquent territory, baseline NIHSS) that might benefit differentially from MT.

We observed moderate statistical heterogeneity for the outcomes of functional efficacy and mortality, and substantial and significant heterogeneity for the outcome of symptomatic intracranial hemorrhage (sICH).

## CONCLUSION

The evidence does not support the routine use of mechanical thrombectomy over standard medical treatment for medium and distal vessel occlusions in terms of functional improvement. Non-significant trends towards increased mortality and sICH with MT were observed, with considerable heterogeneity for sICH. The therapeutic decision should remain individualized, and further research is crucial to define the exact role of MT in this specific population of patients with ischemic stroke.

## Data Availability

All data generated or analyzed during this study are included in this published article and its supplementary information files.

